# Did you miss me? Making the most of digital phenotyping data by imputing missingness with point process models

**DOI:** 10.1101/2025.05.13.25327521

**Authors:** Imogen E. Leaning, Andrea Costanzo, Raj Jagesar, Loran Knol, Sarah Tjeerdsma, Anna Tyborowska, Nessa Ikani, Lianne M Reus, Pieter Jelle Visser, Martien J.H. Kas, Christian F. Beckmann, Henricus G. Ruhé, Andre F. Marquand

## Abstract

**Objectives:** Digital phenotyping has broad clinical potential, providing low-burden objective measures of behaviour as individuals go about their lives. Digital phenotyping goals include the prediction of relapse in mental illness and improved symptom monitoring. However, progress in making clinical inferences from these data is severely challenged by the common occurrence of missing data. We propose a novel method to address this by using non-homogeneous Poisson point process models (PPPMs) to impute missing digital phenotyping data, accounting for their timeseries nature, where smartphone-based activities are modelled as ‘points’.

**Methods:** We demonstrate the use of PPPMs for imputing timeseries data and evaluate their influence on downstream analysis. We evaluate the inclusion of time-varying covariates to model diurnal variations in personalised PPPMs. We validate this model using participants from SMARD (n=26) in a ground truth evaluation, then in PRISM (n=65) and Hersenonderzoek studies (n=283), performing a replication analysis involving hidden Markov models (HMMs).

**Results:** In the ground truth evaluation, PPPMs including ‘hour of the day’ as a covariate, encoded using one-hot encoding, provided the best fit (highest out-of-sample likelihood). Using this imputation method, HMM properties such as daily rhythms were preserved and we successfully replicated findings from our prior work.

**Discussion:** Personalised PPPMs using covariates provide tailored simulations of behaviour that can be used for imputation in behavioural time series.

**Conclusion:** PPPMs using covariates are a promising imputation tool that may contribute to improved utility of digital phenotyping.

## Introduction

Digital phenotyping is a highly promising technique to facilitate the quantification of behavioural or physiological markers from digital measures, enabling the passive monitoring of patients in a naturalistic environment. Key clinical goals include the objective measurement of behaviour and prediction of relapse in mental illness. An emerging body of digital phenotyping investigations have produced promising results, including demonstrating correlations between digital phenotyping features and depression symptoms[1, 2], and age and diurnal effects[3]. Several apps exist to record digital phenotyping data on smartphones, including ‘Behapp’[4] and ‘RADAR-base pRMT’[5]. However, across studies and platforms the theme of missing data has emerged as a major hurdle[6], hindering clinical implementation.

Missing data may arise due to user behaviour, e.g. switching on flight mode, or technical issues, such as changes in app permissions, battery management software or sensor problems. In a systematic review of digital phenotyping in major depression (MDD)[7], 20/24 articles referred to missing or absent data. To handle missingness, samples were often excluded from analysis, with ten studies using a threshold for exclusion and three studies excluding samples if any variable was missing. However, this can introduce bias[8] and lead to the loss of large volumes of data[9]. In this review, only five studies mentioned carrying out some kind of imputation, including using the mean from preceding data[10], applying a resampling method to GPS data[11], using linear interpolation on Bluetooth data[12] and imputing missing records using similar records[13].

Imputation is useful as it increases the number of usable samples, therefore increasing the power of the statistical analyses and broadening the number of methods available, as researchers are not limited to methods that can handle missingness. However, timeseries imputation has additional challenges due to the temporal dependency between timepoints. Popular approaches for imputing time series include simple methods such as using the mean/mode or last-observation-carried-forward, or more advanced methods that take advantage of relationships between variables (e.g. Multivariate Imputation by Chained Equations (MICE)[14]). Mean/mode or last-observation-carried-forward methods result in the same value being used for imputation, which may warp the data distribution. Additionally, missingness may occur across all variables at the same timepoints due to app malfunctions, meaning that methods such as MICE are unsuitable as when missingness occurs there are no available points of information in any variables.

Within smartphone-based digital phenotyping, there have been limited direct investigations of appropriate methods for imputation. One study used Gaussian process models to impute GPS data[15], and another study applied regression models to impute social media features[16]. Many of the direct imputation investigations have occurred within wearables research, in particular for accelerometer data. Methods include multiple imputation[17, 18], neural networks[19], and k-nearest neighbors[8, 20]. Accelerometer and GPS data have high temporal resolution, contrasting with sparser digital phenotyping channels, such as app usage. Additionally, imputation methods that rely on other datapoints being present cannot be used in cases where missingness occurs across all channels concurrently.

To address these problems, we aimed to investigate Poisson point process models (PPPMs) as a method to impute missing digital phenotyping data occurring in activity-based channels. PPPMs model the probability of ‘points’, in our case activities, occurring. We evaluated the imputation of phone usage activities collected using the Behapp platform[21] (Figure 1), although this approach is suitable for any platform. First, we investigated PPPM performance against a ground truth from participants in the Smartphone based Monitoring and cognition modification Against Recurrence of Depression (SMARD) dataset[22]. Models were evaluated both in- and out-of-sample, and imputation was carried out by simulating point processes using the PPPMs (Figure 1d). We evaluated the impact of imputation on downstream analysis by comparing hidden Markov models (HMMs)[23] trained separately on imputed data and ground truth data (Figure 1e). Next, we validated the use of PPPMs for imputation in a real missingness case from the Psychiatric Ratings using Intermediate Stratified Markers (PRISM)[24] and Hersenonderzoek[25] datasets (Figure 1f). Specifically, we replicated the analysis in Leaning et al.[23] using imputed data (Figure 1g), to evaluate whether imputation would impact this paper’s findings (Figure 1h).

**Figure 1.**
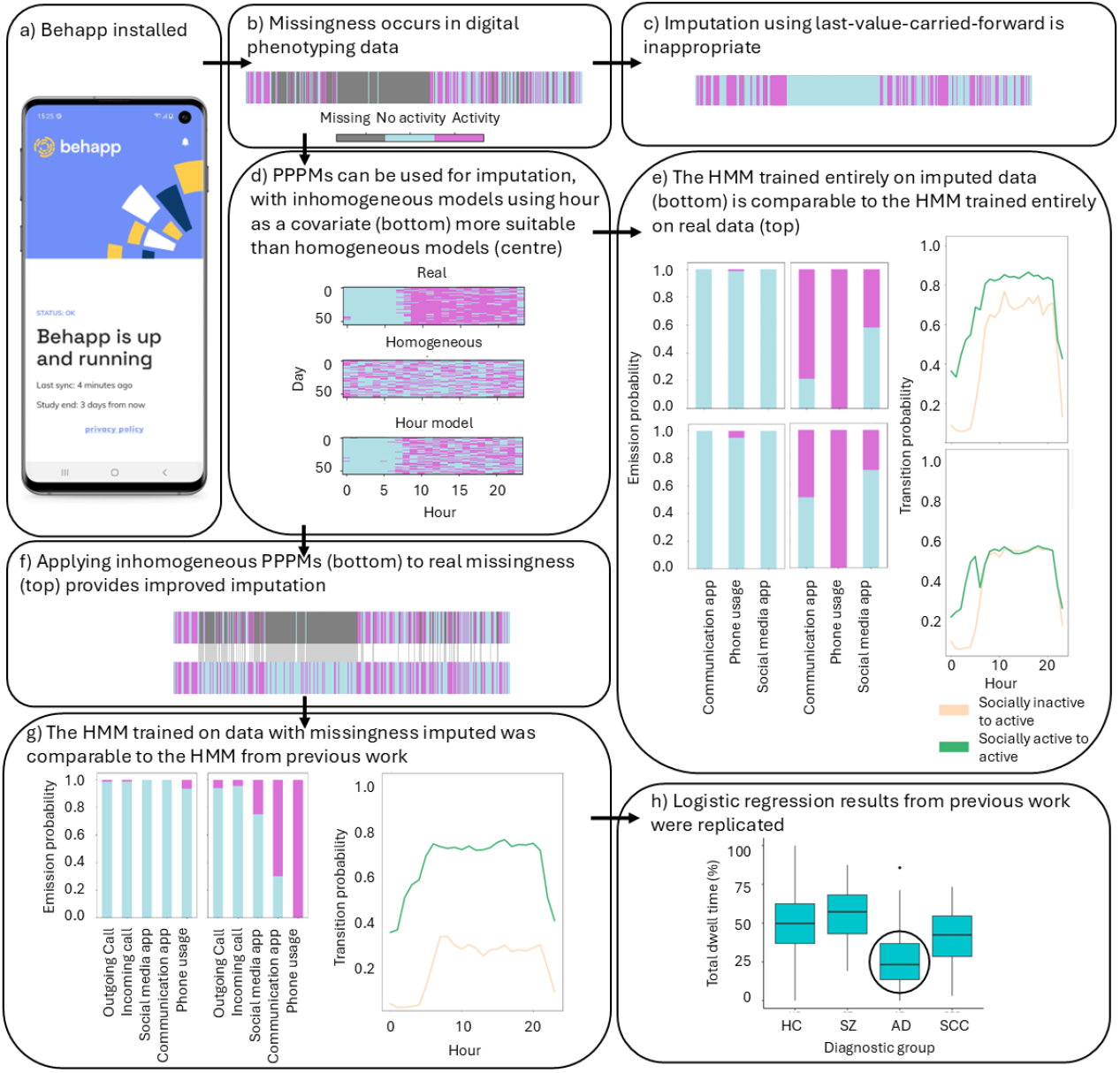
Overview of the imputation method. a) Behapp was installed on participants’ smartphones, however b) missingness is common. c) Methods like last-value-carried-forward provide poor imputation. d) Poisson point process models (PPPMs) are more suitable. e) When compared to a hidden Markov model (HMM) trained on real data, a HMM trained on imputed data remained similar. f) PPPM-based imputation is more realistic than the imputation in c). g) A HMM trained on data with missingness imputed using PPPMs remained very similar to the HMM from the original study, with h) previous results replicated.

## Methods

### Datasets

Participants from the SMARD dataset[22] in the Netherlands were used for the first part of this study. SMARD aims to investigate depression recurrence in recurrent MDD. Participants were between 18-65 years, with a diagnosis of MDD and classified as being in remission. Information on inclusion criteria is provided in Ikani et al.[22]. Behapp collection lasted for 1.5 years/participant. A subset of participants who had excellent data availability (≤1% missingness) were selected, to ensure sufficient ground truth data were available for evaluation. Data collection took place between August 2019–March 2023.

The second part of the study used data from the PRISM[26] and Hersenonderzoek[25] studies, which collected Behapp data for 42 days/participant. Study details are provided in the original analysis[23]. Data collection took place between August 2017-May 2019 for PRISM and March 2018-January 2020 for Hersenonderzoek. Both datasets contained participants with probable Alzheimer’s disease (AD) and age-matched healthy controls (HCs), PRISM contained participants with schizophrenia (SZ) and Hersenonderzoek contained participants with subjective cognitive complaints (SCC). PRISM investigates social withdrawal in SZ and probable AD[27, 28]. Recruitment took place in the Netherlands and Spain. Participants with SZ were between 18-45 years, and participants with probable AD were between 50-80 years. In Hersenonderzoek, participants in the Netherlands with age 45+ years self-reported probable AD, or an absence of neurological or psychiatric diseases with or without memory complaints (SCC and HCs respectively)[25].

### Behapp acquisition and smartphone channels

‘Behapp’ was installed on each participant’s smartphone[21]. Social media app, communication app and overall phone usage activities were recorded for all studies. Outgoing/incoming calls were additionally recorded for PRISM and Hersenonderzoek. The start and end times for each activity were recorded. Activities were split into hourly bins indicating whether an activity was carried out each hour, with missing hours identified (see the supplement and Leaning et al.[23] for details). Missingness occurred across all channels concurrently.

### PPPMs

A Poisson distribution models the probability of a point occurring within a given domain, e.g. the probability of app use across time. We treated homogeneous PPPMs, which model a constant probability of a point occurring, as our baseline. We evaluated non-homogeneous PPPMs against this baseline, where the probability varies according to the values of covariates. We used the ‘NHPoisson’ R package (version 3.3)[29] for training, covariate selection and simulation. Full details are provided in the supplement.

### Model development: Ground truth-based imputation investigations

For each participant, we fit homogeneous and non-homogeneous PPPMs where ‘hour of the day’ and ‘day of the week’ were used as covariates, in order to model diurnal and ultradian rhythms. We investigated two covariate encoding methods: one-hot encoding and sine-cosine transformation (see supplement). We used 10-fold cross-validation to investigate PPPM performance, where for each participant at each iteration their withheld fold was considered ‘missing’. PPPMs were trained for each available activity channel (overall phone usage, communication app usage, social media app usage).

For each channel, likelihood ratio tests (LRTs) were run for each trained model, with comparisons made between the homogeneous model and models that included one of: one-hot encoded hour, sine-cosine transformed hour, one-hot encoded day and sine-cosine transformed day. The *p-*value from the LRT was used to assess whether the covariate model provided a significantly better fit (*p*<.05). For participants where the day covariate was significant for ≥1 fold (considering each encoding method), additional models were trained that included both day and hour, with LRTs carried out for this model compared to the hour-only model, to verify whether the day was contributing significantly more information than the hour alone.

The accuracy of the trained models was evaluated by calculating the out-of-sample Poisson log-likelihood of the datapoints in each of the withheld folds, given the corresponding model trained on the other nine folds, and calculating the mean across folds per participant for each PPPM type. The trained models were used to simulate events occurring during the ‘missing periods’, by simulating point processes during a time period matching the withheld fold in terms of length and covariate values using NHPoisson’s ‘simNHP’ function. The simulated time series could be compared to the real values for this fold.

After selecting the best performing PPPM type, the simulated time series for each channel were concatenated and used to train HMMs following the method in Leaning et al.[23] using the ‘depmixS4’ package[30], with the one-hot encoded hour of the day included as a covariate on the HMM transition probabilities, to investigate the effect of imputation on downstream analysis. We considered ‘unconstrained’ and ‘constrained’ imputation cases (see supplement for details) and compared these HMMs to a HMM trained on the ground truth data.

### Clinical validation: Imputation application to real missingness

Next, we applied the selected imputation method to data from the PRISM[26] and Hersenonderzoek[25] studies used in a prior study (where missingness was not imputed), replicating the key findings[23]. Where missingness occurred, we inserted the values simulated by each personalised PPPM (Figure 1f).

We then trained a HMM as in Leaning et al.[23]. We calculated the ‘total dwell time’ per participant, i.e. the percentage of time spent in the identified ‘socially active’ hidden state. We compared total dwell times between each diagnostic group (SZ, AD, SCC) and HCs using logistic regression. We present FDR-corrected *P* values, correcting for 3 tests, with results considered significant at *P*<.05.

## Results

### Sample statistics

Demographics are provided in Table 1. SMARD participants had experienced a mean of 4.4(±2.5) depressive episodes. Their mean time series length was 213(±214) days. For PRISM, the mean length was 34(±13) days. For Hersenonderzoek, the mean length was 35(±12) days. Further details are provided in Leaning et al.[23].

**Table 1.** Demographics across diagnostic groups.

| Diagnostic group | Number | Dataset | Age (Mean (SD)) | Sex | Country |
| --- | --- | --- | --- | --- | --- |
| Major depression | 26 | SMARD | 51(±14) | F=22; M=4 | NL=26 |
| Healthy control | 247 | PRISM=28;<br>Hersenonderzoek=219 | 59 (13) | F=140; M=107 | NL=234; ES=13 |
| Schizophrenia | 18 | PRISM | 31 (6) | F=7; M=11 | NL=12; ES=6 |
| Alzheimer’s disease | 26 | PRISM=19;<br>Hersenonderzoek=7 | 67 (7) | F=10; M=16 | NL=18; ES=8 |
| Subjective cognitive complaints | 57 | Hersenonderzoek | 61 (7) | F=36; M=21 | NL=57 |

*SD: standard deviation; F: female, M: male; NL: the Netherlands, ES: Spain.*

### Ground truth-based imputation investigations

The percentage of models for which each covariate was significant in LRTs are provided in supplementary Table S1. For almost all models, hour was significant. The day covariate was less commonly significant, with the maximum percentage of models for which this was significant at 30% (communication app channel). For models containing both day and hour, day generally remained significant (supplementary Table S2).

The out-of-sample Poisson log-likelihoods are shown in Figure 2 (with the numerical values in supplementary Tables S3-S5). The log-likelihoods for PPPMs including hour were higher than for homogeneous PPPMs, indicating a better fit. PPPMs including one-hot encoded hour were also generally higher than sine-cosine transformation. When both day and hour were included, the log-likelihood rarely showed an improvement upon models with only hour: models including both covariates provided the highest log-likelihood for 0/7 participants for phone usage, 5/13 participants for communication app (with 2 of these negligibly higher), and 2/14 participants for social media app (with 1 of these negligibly higher). We therefore decided to use PPPMs including one-hot encoded hour as our imputation approach for the remaining analyses.

**Figure 2.**
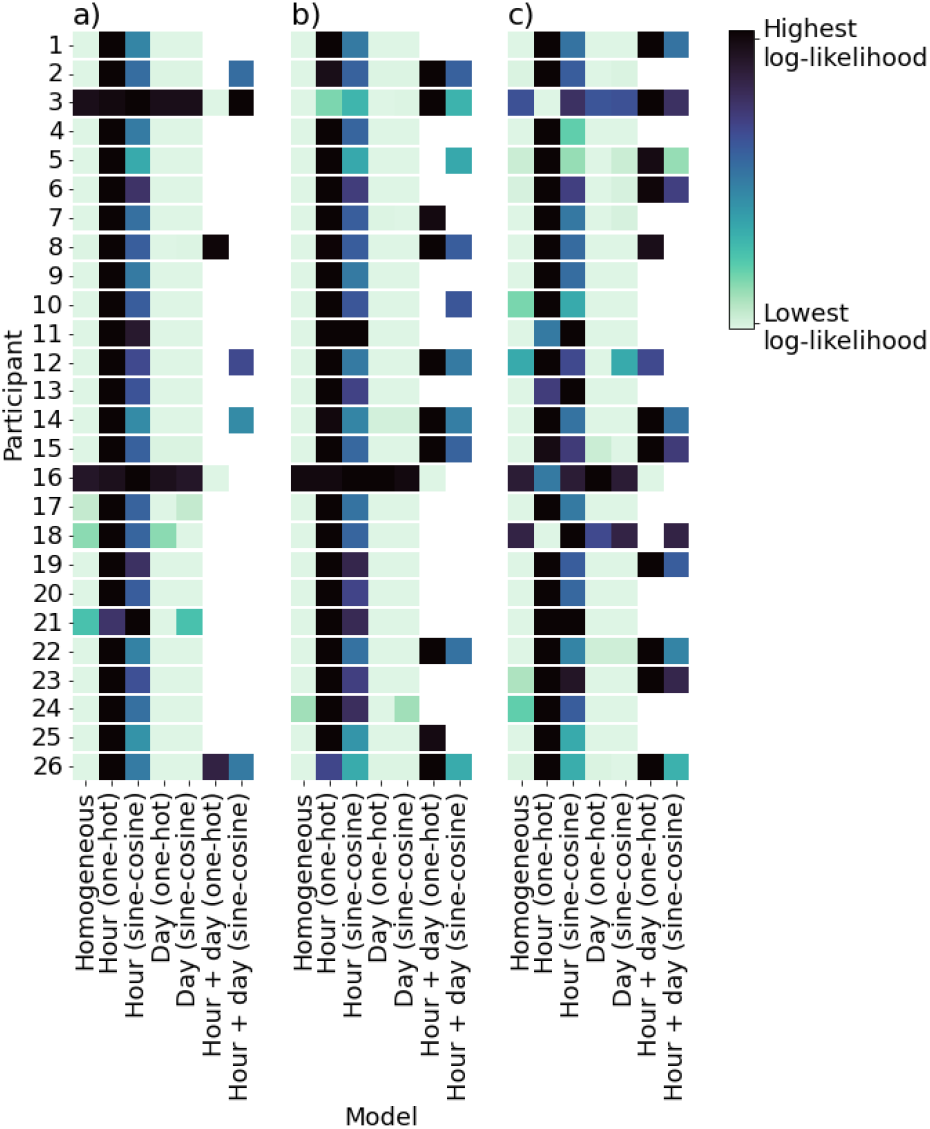
Mean out-of-sample log-likelihoods for a) overall phone usage, b) communication and c) social media apps. Each row corresponds to one participant. The darkest tile gives the highest mean log-likelihood for a participant.

We present simulated visualisations alongside real data in Figure 3 and supplemental Figures S1-S2. Figures 3a and 3b display participants with clear diurnal rhythms, and 3c shows a participant with elevated nighttime activity. PPPMs including hour provided improved imputation when compared to homogeneous PPPMs, capturing personal diurnal behaviours.

**Figure 3.**
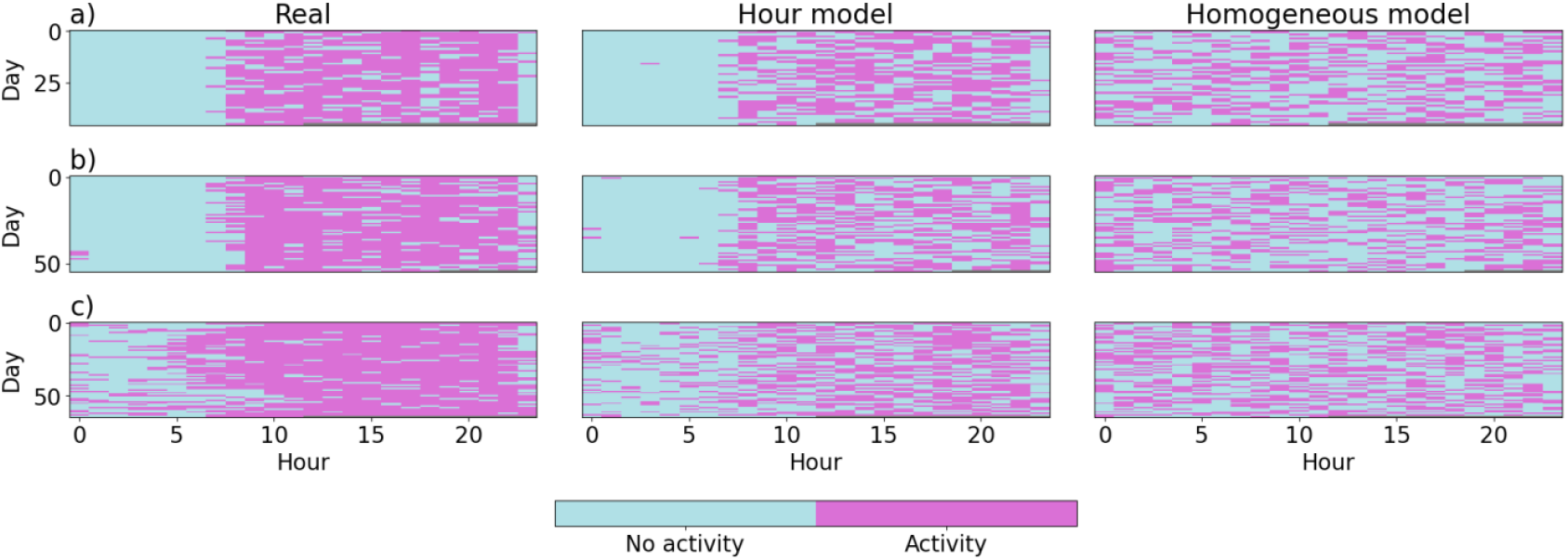
Real versus imputed time series for the phone usage channel for three participants (a), b) and c)). Each row corresponds to a single fold from each participant. The left column shows the fold’s original time series, the centre shows the simulation provided by the PPPM using one-hot encoded hour, and the right shows the corresponding homogeneous PPPM. For each participant, the hour model captured individual behaviour more accurately than the homogeneous model.

The simulated SMARD data was concatenated and used to train HMMs which were compared to an HMM trained entirely on the ground truth (Figure 1e). The ground truth HMM has one state with very low probabilities of activity across channels, and another state with a very high probability of activity in the phone usage channel, high probability of communication app usage and lower probability of social media app usage (supplemental Figure S3a). The HMM trained on unconstrained imputed time series (supplemental Figure S3b) also shows low activity and high activity states, however the high activity state here gives lower probabilities than the ground truth, with phone usage still the most probable channel. Once the imputed time series is constrained (supplemental Figure S3c), the probability of phone usage is more comparable to the ground truth HMM.

For all HMMs, the probability of transitioning into the high activity state, from either state, rapidly increases during the morning, is high throughout the day and decreases in the evening, though varies in magnitude (supplemental Figure S4). For both HMMs trained on imputed data, there was a 1.0 probability of starting the time series in the inactive state, whereas for the HMM trained on the real data, this probability was 0.44.

### Imputation application to real missingness

After applying the imputation method to missingness in PRISM and Hersenonderzoek (Figure 1f), the percentage of missing datapoints decreased from 30% to 4%, with the 4% missingness from participants where imputation was not reasonable. Example real time series and their corresponding simulated time series are shown in Figure 4. We trained a 2-state HMM using the updated time series (with missingness imputed) (Figure 1g).

**Figure 4.**
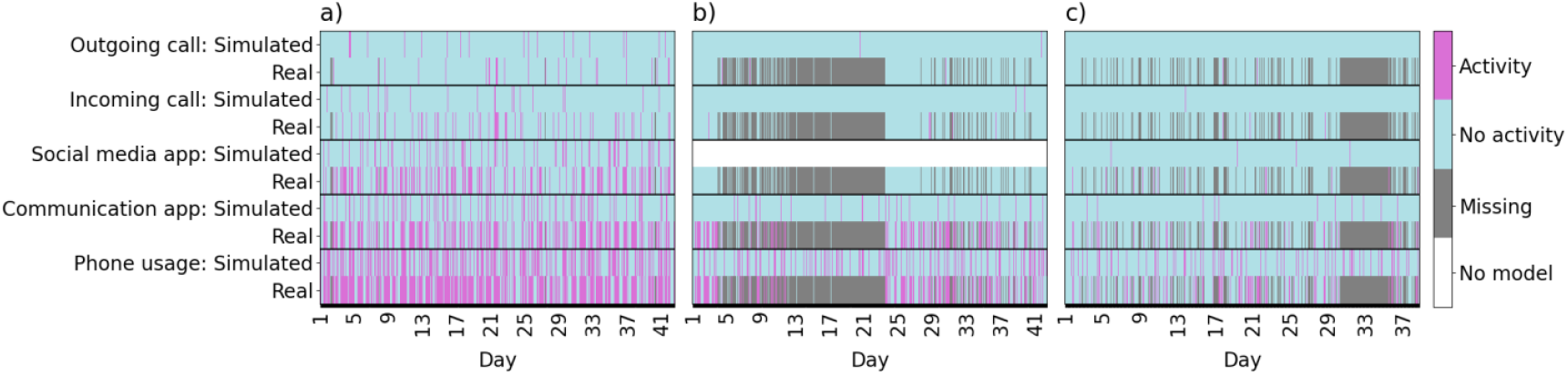
Example real time series from three participants with their corresponding simulations, used to impute the missing periods. a) A participant with very low missingness. The simulated time series show similar patterns, most obvious in the phone usage channel, but underestimate the number of points; b) a participant with notable missingness; c) a participant with moderate missingness.

The emission probabilities from the HMM containing imputation are extremely similar to those from the original HMM, reflecting ‘socially active’ and ‘socially inactive’ states (see supplemental Figure S5). The transition probabilities show the same pattern of increased probability of transitioning into the socially active state during the daytime (see supplemental Figure S6).

We then successfully replicated the prior logistic regression results (Figure 1h), with the time spent socially active significantly lower in the AD (FDR-corrected *P*<.001) and SCC (FDR-corrected *P*=.01) groups relative to HCs (see supplemental Figure S7 and Tables S6-S7). These results were seen for both constrained and unconstrained imputation.

## Discussion

In this work, we demonstrated the utility of PPPMs as a method for imputing missing smartphone timeseries activity data. In our ground-truth investigations, we investigated homogeneous PPPMs and PPPMs that included covariates. We identified that the PPPMs including one-hot encoded hour of the day generally provided the best models out-of-sample, suggesting that accounting for diurnal variation is important for imputation. HMMs trained on imputed data learnt comparable model parameters to HMMs trained on original data, preserving properties of the original data.

We investigated the influence of our imputation method on a prior analysis that used HMMs to investigate group differences between various clinical populations and HCs. When imputation was applied, the updated HMM was highly comparable to the original HMM. We subsequently demonstrated significantly lower time spent socially active for participants with AD and SCC compared to HCs, consistent with the previous study.

We selected the best performing PPPM across participants for imputation. However, future work could carry out individualised covariate selection. Additional covariates that may affect behaviour, e.g. weather, could be explored. The suitability of using PPPMs to predict activity points with an associated continuous value could be investigated, e.g. using marked PPPMs. In our ground truth HMM investigations, we compared real data to an extreme missingness case where all data was ‘imputed’. Future work should seek to investigate the limits to how much data can realistically be imputed using PPPMs. Non-homogeneous PPPM could be applied to additional data types, such as ecological momentary assessment data. The PPPM method could also be used to infer whether a person has diurnal and/or weekly rhythms as its own question of interest.

We treated a homogeneous PPPM as our baseline, however, to our knowledge, this is also a novel imputation method for digital phenotyping. Several other methods had obvious limitations (Figure 1c), however it would have been preferable to have been able to use a more established baseline. Our approach of applying PPPMs to each channel does not account for covariation between different channels. Future research could seek to investigate multivariate imputation using hierarchical or multivariate PPPMs. Whilst our datasets were prone to chunks of missingness, in some participants missingness occurs periodically (see Figure 4c). In these cases, further refinement may be needed, such as including parameter values from similar participants without periodic missingness. Additional user behaviours (e.g. phone switch-offs) may explain this missingness and so could be recorded. As our datasets include clinical populations, it is possible that their time series include behavioural changes that can be attributed to symptoms. For our initial investigations we wanted to train with as much data as possible, however it could be evaluated whether to use all data for imputation or to select periods meeting certain criteria.

## Conclusions

Digital phenotyping has high clinical potential but is prone to missingness. We investigated non-homogeneous PPPMs for imputing missing activity in time series. Non-homogeneous PPPMs using hour of the day encoded using one-hot encoding provided the highest out-of-sample Poisson likelihood, so were selected as our imputation method. We replicated key results from a previous study, with missingness imputed using the selected PPPM type. Using non-homogeneous PPPMs for imputation is a promising avenue for improving the analysis and utility of digital phenotyping.

## Contributors

IEL contributed to study conceptualisation, formal analysis, methodology, software, visualisation and writing – original draft. AC and RJ contributed to Behapp software, PRISM data curation, investigation and writing – review & editing. LK contributed to study conceptualisation and writing – review & editing. ST, AT and NI carried out SMARD data curation and writing – review & editing. LMR and PJV carried out Hersenonderzoek data curation and writing – review & editing. MJHK contributed to study conceptualisation, funding acquisition, project administration and writing – review & editing. CFB contributed to study conceptualisation, supervision and writing – review & editing. HGR contributed to study conceptualisation, funding acquisition, supervision and writing – review & editing. AFM contributed to study conceptualisation, funding acquisition, methodology, supervision and writing – review & editing.

## Competing interests

CFB is a director of SBGNeuro. HGR received grants from the Hersenstichting, ZonMw, the Dutch Ministry of Health and an unrestricted educational grant from Janssen. In addition he received speaking fees from Lundbeck, Janssen, Benecke and Prelum; all outside the current work. All other authors declare no conflicts of interest.

## Funding

This study was funded by the European Research Council (consolidator grant 101001118). The SMARD study was funded by the Hersenstichting (Dutch Brain Foundation) (HA2015.01.07). The Dutch Brain Research Registry (Hersenonderzoek) is supported by ZonMw‐Memorabel (project no 73305095003), Alzheimer Nederland, Amsterdam Neuroscience, and Hersenstichting. The PRISM project has received funding from the Innovative Medicines Initiative 2 Joint Undertaking under grant agreement 115916. This Joint Undertaking receives support from the European Union’s Horizon 2020 research and innovation programme and EFPIA. This study reflects only the authors’ view and the European Commission is not responsible for any use that may be made of the information it contains.

## Data availability

The datasets analysed during the current study are available from the corresponding author on reasonable request.

## Code Availability

The imputation method script is available in https://github.com/predictive-clinicalneuroscience/Imputation_Digital_Phenotyping, and the scripts related to the HMM part of the analysis are available in https://github.com/predictive-clinicalneuroscience/HMM_Digital_Phenotyping

## Ethics Approval

SMARD was approved by the Radboudumc ethics review board (2016-3009; NL60033.091.16). PRISM was approved by the Ethical Review Board University Medical Centre of Utrecht (17-021/D) for the Dutch research centres, and the Comité Ético de Investigación Clínica Hospital General Universitario Gregorio Marañón (59359) for the Spanish research centres. Hersenonderzoek was approved by the Ethical Review Board VU University Medical Centre (2017.254). All participants provided informed consent before participating.

## Notes

### Competing Interest Statement

Christian Beckmann is a director of SBGNeuro. Henricus Ruhé received grants from the Hersenstichting, ZonMw, the Dutch Ministry of Health and an unrestricted educational grant from Janssen. In addition he received speaking fees from Lundbeck, Janssen, Benecke and Prelum; all outside the current work. All other authors declare no conflicts of interest.

### Author Declarations

SMARD: The ethics committee of the Radboudumc (Radboudumc ethics review board (2016-3009; NL60033.091.16)) gave ethical approval for this work. PRISM: The ethics committee of the University Medical Centre of Utrecht (Ethical Review Board University Medical Centre of Utrecht (17-021/D)) gave ethical approval for this work at the involved Dutch research centres. The ethics committee of Hospital General Universitario Gregorio Marañón (Comité Ético de Investigación Clínica Hospital General Universitario Gregorio Marañón (59359)) gave ethical approval for this work at the involved Spanish research centres. Hersenonderzoek: The ethics committee of VU University Medical Centre (Ethical Review Board VU University Medical Centre (2017.254)) gave ethical approval for this work.

### Summary of Updates

The manuscript text has been streamlined to reduce length. Important methodological details are retained in the main manuscript, and further details have been moved to the supplement. The transition probability figures have also been corrected.

