## Supplementary materials for "Did you miss me? Making the most of digital phenotyping data by imputing missingness with point process models"

Further methodological details are provided in this supplement, as well as additional figures and tables.

#### Additional details on measures

We identified hourly missingness by using the expected sampling frequency (>1/hour) of sensors including GPS and WiFi, which do not depend on active phone usage; if these sensors did not provide any datapoints for an hour this hour was considered to be missing. Consequently, any missing points occurred at the same time points across all channels. Whilst some missingness appeared periodically, data were prone to seemingly random chunks of missingness (see Figure 4 in the main text).

Each app used by a participant is classified according to its label in the Google Play Store. Whilst Behapp also collects GPS data, we focused on phone usage-related data as event-based channels are more suitable to be modelled as points. The first day of each participant's data was excluded in case of onboarding effects.

#### Poisson point process model details

Homogeneous PPPMs were chosen as our baseline method, as many other common imputation models would be nonsensical (see Figure 1c in the main text). Training was carried out using the 'fitPP' function. During model fitting, the negative of the Poisson log-likelihood was minimised using NHPoisson's default function, 'nlminb'.
The Poisson log-likelihood of a series of points given the learnt model is given by the equation:

$$LL(\boldsymbol{\beta}; (t_i)_{i=1}^{n}) = -\sum_{t=1}^{T} \lambda(t; \boldsymbol{\beta}) + \sum_{i=1}^{n} \log \lambda(t_i; \boldsymbol{\beta}) \quad (1)$$

where

$$\lambda(t; \boldsymbol{\beta}) = \exp(\mathbf{X}^{\top}(t)\boldsymbol{\beta}). \quad (2)$$

$\boldsymbol{\beta}$ is the vector of model parameters from the trained model, $t$ is each individual timepoint, $n$ is the total number of points with activity within the given time series, $\mathbf{X}^{\top}(t)$ is the vector of time-varying covariates at time $t$ and $T$ is the length of the time series. The first part of the equation therefore is calculated across the whole time series, and the second part of the equation is calculated for the timepoints with activity.

We investigated two different covariate encoding methods. One-hot encoding involves converting each covariate value into an array of length n (number of possible covariate values) minus 1, i.e. length 6 for day of the week and 23 for hour of the day. An array of zeros is used where for each covariate value one of the positions is set to 1, with the exception of one value whose corresponding array is entirely zeros. Sine-cosine transformation instead requires two dimensions to encode each value, $v$, using sine and cosine functions:

$$x = \sin\left(\frac{v}{p} \times 2\pi\right) \quad (3)$$

and

$$y = \cos\left(\frac{v}{p} \times 2\pi\right), \quad (4)$$

where $p$ is the relevant period (e.g. $p = 24$ for the hour covariate).

We carried out validation using 10-fold cross-validation. Each participant's time series was split into ten equal-length consecutive segments. For each iteration, 9 folds of a participant's time series were used to train each PPPM and the remaining fold was considered to be 'missing', with the actual values from each 'missing' fold used as the ground truth. This then meant that for each activity channel, 10 models were trained per participant for each PPPM type. To simulate time series using PPPMs, Equation 2 was used to provide the value of $\lambda$ used by the 'simNHP' function.

#### Hidden Markov model details

To investigate the effect of PPPM-based imputation on downstream analysis, we evaluated its impact on a Hidden Markov Model (HMM). In part 1, we trained a HMM on the original SMARD data, taking this to be the ground truth HMM, and compared this to HMMs trained on data simulated by the trained PPPMs. Two HMMs concerning imputed data were considered: an HMM trained on the original imputed time series for each channel, and an HMM trained on imputed time series that were constrained by the imputed values in the phone usage channel. This constraint was imposed due to the nature of the channels, as it is not possible to have communication or social media app usage without phone usage. For the timepoints where there was no phone usage activity, we set the values of these channels to no activity. The latter model provides the more plausible behavioural pattern, although this of course reduces the number of activity points. We compared each of these HMMs to the HMM trained on the corresponding 'real' data to investigate whether the HMM properties, i.e. the emission, transition and starting probabilities, were preserved.

In part 2, we carried out a replication analysis. We repeated the key parts of the prior analysis[1], where a HMM was trained on the Behapp channels: phone usage, communication app, social media app, incoming and outgoing calls. A measure derived from the HMM, the 'total dwell time', was compared between the different diagnostic groups. Our key finding from this paper was that participants with AD and SCC had lower total dwell time than HCs (i.e. they spent less time in the socially active hidden state). We sought to replicate the hidden states identified in Leaning et al.[1], training the HMM on an updated time series where missingness was imputed using PPPMs, and to evaluate whether we would still identify these significant group differences.

In the current study, we used the available data per participant to train an individualised non-homogeneous PPPM with one-hot encoded hour as a covariate. For participants whose model failed to converge, the 'optim' function with the 'BFGS' method was used in place of the default function. For participants who had a channel with less than 3 activity points, we did not carry out imputation for that channel due to the limited amount of data available for training. This threshold was arbitrarily selected as for some channels datapoints could be very sparse. Additionally, if a participant had more than 85% of their time series missing no imputation was carried out as missingness was deemed too high for a representative model to be learnt. This threshold required almost a week's worth of data per participant.

We trained a two-state HMM on the completed time series (with missingness now imputed), as per the method in Leaning et al.[1]. We chose to train the model on all participants rather than the training split used in the main text of this paper so that we could investigate the effect of imputation on the HMM for as many participants as possible, and therefore compared our results to the supplementary results of this paper that did not include a training/validation split. Note that the results from this original supplementary two-state model were consistent with the results from the original main reported model. We verified our findings for both constrained and unconstrained imputation approaches, as done in part 1 of the study.

### Figures

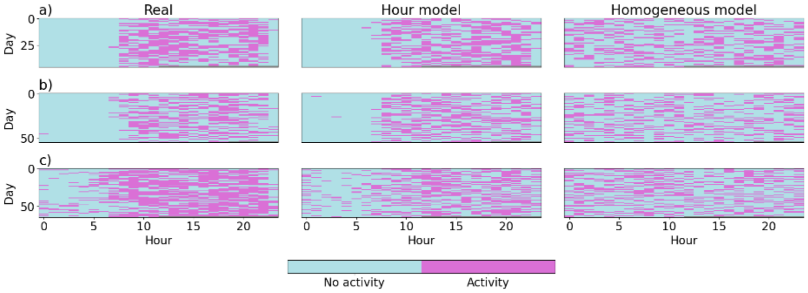


Figure S1. Real versus imputed time series for the communication app channel for three participants (a), b) and c)). Each row corresponds to a single fold from each participant from their cross-validation. The left column shows the fold's original time series, the centre column shows the simulation provided by the PPPM using one-hot encoded hour, and the right column shows the corresponding homogeneous PPPM.

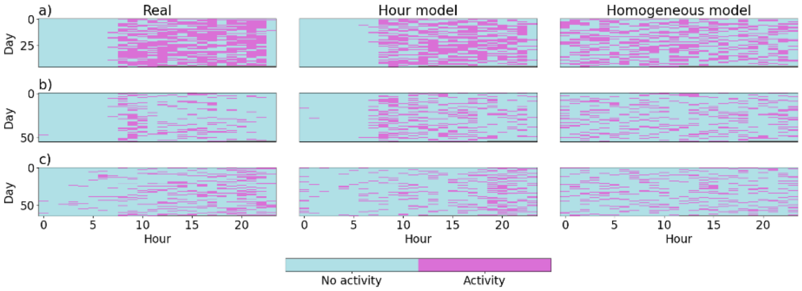


Figure S2. Real versus imputed time series for the social media app channel for three participants (a), b) and c)). Each row corresponds to a single fold from each participant from their cross-validation. The left column shows the fold's original time series, the centre column shows the simulation provided by the PPPM using one-hot encoded hour, and the right column shows the corresponding homogeneous PPPM.

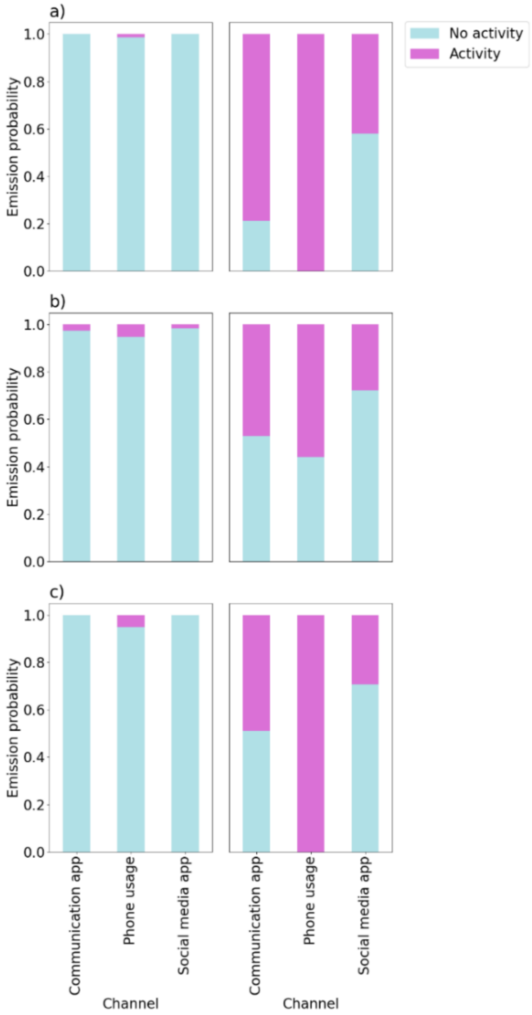


Figure S3. Emission probabilities from the different HMMs. The HMMs were trained entirely on a) real data; b) imputed data, where channel values were unconstrained by phone usage; c) entirely on imputed data, where channel values were constrained by phone usage. Each HMM has a low activity state (lefthand side) and a state with higher probabilities of activity (righthand side). The high activity states are referred to as 'socially active' and the low activity states as 'socially inactive', for consistency with the second part of this study.

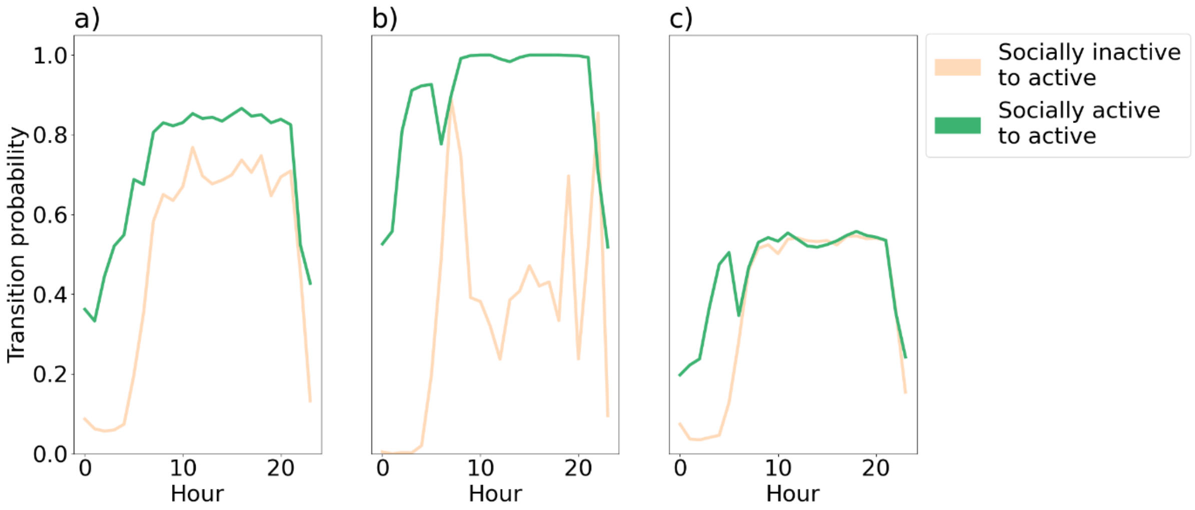

Figure S4. Transition probabilities between hidden states from the different HMMs. The HMMS were trained entirely on a) real data; b) imputed data, where channel values were unconstrained by phone usage; c) imputed data, where channel values were constrained by phone usage. The high activity states are referred to as ‘socially active’ and the low activity states as ‘socially inactive’.

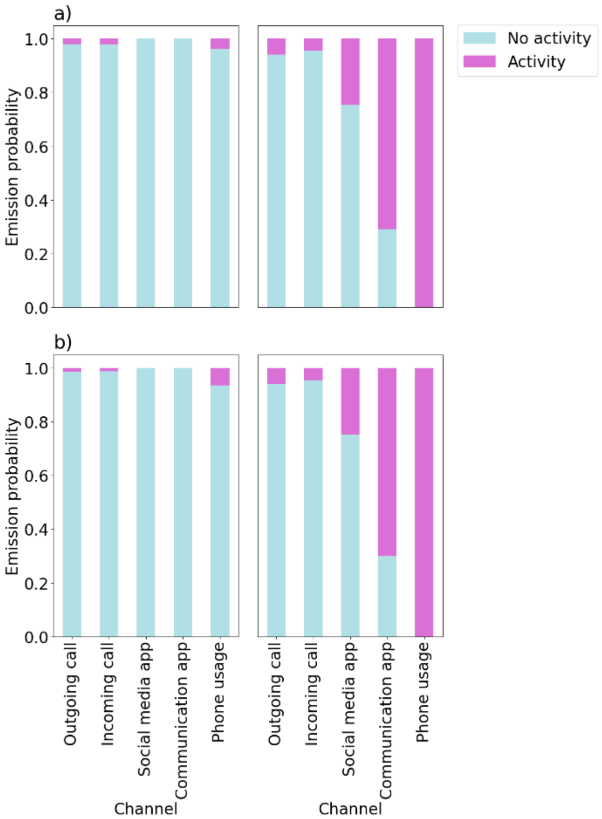


Figure S5. Emission probabilities for the original HMM and the replicated HMM. a) The two state HMM from the original paper, trained using all participants and b) the HMM trained in the same way except with missing data imputed (and constrained based on phone usage). The states on the lefthand side of a) and b) show a very low probability of activity, whereas the righthand side shows higher probabilities of phone and app usage. Differences between a) and b) are very small.

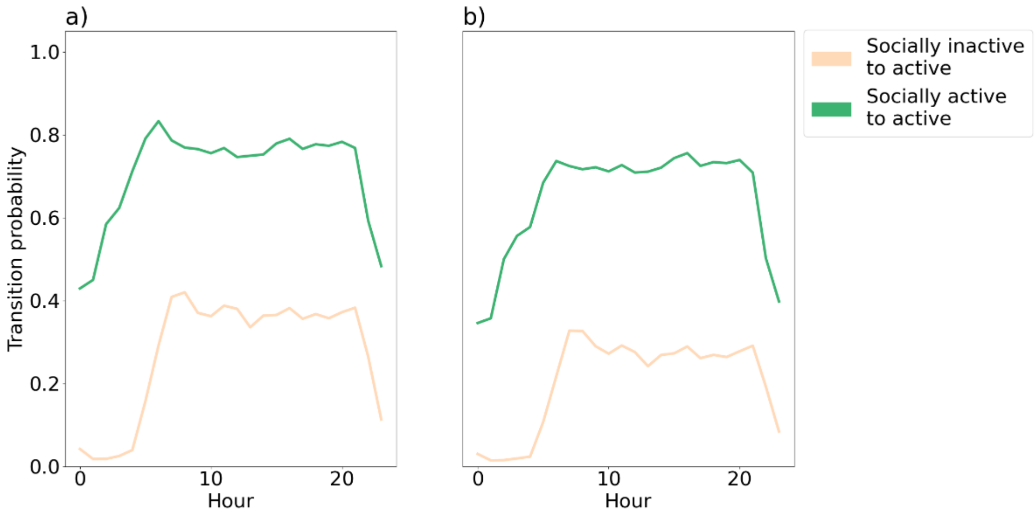

Figure S6. Transition probabilities for the original HMM and the replicated HMM. a) The two-state HMM from the original paper, trained using all participants and b) the HMM trained in the same way except with missing data imputed (and constrained based on phone usage). For both a) and b), the probability of transitioning into the socially active state from either state is higher during the daytime than the nighttime. Differences between a) and b) are small. 0: midnight.

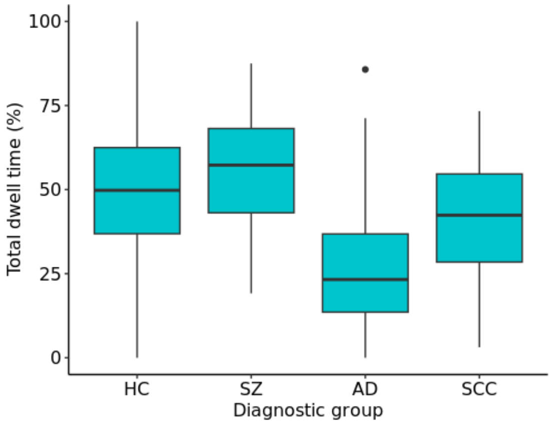


Figure S7. A box plot of the total dwell times per participant for the different diagnostic groups, using the HMM that had imputation constrained based on phone usage. There is a significant difference between the HC and AD groups, and the HC and SCC groups. HC: healthy control, SZ: schizophrenia, AD: Alzheimer's disease, SCC: subjective cognitive complaints.

### Tables

Table S1. Percentage of PPPMs for which each covariate is significant according to likelihood ratio tests (total number of models per row=260). 14/21 of the models that failed to converge were from 1 participant. n=number.

| Channel | Transform | Significant day models (% (n)) | Significant hour models (% (n)) | Day models that failed to converge (% (n)) | Hour models that failed to converge (% (n)) |
|---|---|---|---|---|---|
| Phone usage | One-hot | 8 (21) | 99 (258) | 1 (3) | 1 (2) |
| Phone usage | Sine-cosine | 13 (34) | 100 (260) | 0 | 0 |
| Communication app | One-hot | 30 (79) | 99 (257) | 1 (3) | 1 (3) |
| Communication app | Sine-cosine | 23 (61) | 100 (260) | 0 | 0 |
| Social media app | One-hot | 27 (71) | 98 (254) | 2 (4) | 2 (5) |
| Social media app | Sine-cosine | 16 (42) | 98 (255) | 0.4 (1) | 0 |

Table S2. Likelihood ratio test results comparing models with both day and hour to models solely with hour (for participants where the day covariate was significant).

| Channel | Transform | Significant combined day and hour models/significant hour models (%) |
|---|---|---|
| Phone usage | One-hot | 9/21 (43) |
| Phone usage | Sine-cosine | 33/34 (97) |
| Communication app | One-hot | 63/79 (80) |
| Communication app | Sine-cosine | 60/61 (98) |
| Social media app | One-hot | 60/71 (85) |
| Social media app | Sine-cosine | 42/42 (100) |

Table S3. Mean out-of-sample log-likelihoods for the overall phone usage channel. The highest likelihoods are highlighted.

| Participant | Homogeneous | Hour (one-hot) | Hour (sine-cosine) | Day (one-hot) | Day (sine-cosine) | Hour + day (one-hot) | Hour + day (sine-cosine) |
|---|---|---|---|---|---|---|---|
| 1 | -776 | -683 | -733 | -776 | -776 | | |
| 2 | -1126 | -898 | -1002 | -1125 | -1125 | | -1002 |
| 3 | -1334 | -1298 | -1262 | -1334 | -1333 | -2495 | -1262 |
| 4 | -117 | -111 | -114 | -117 | -117 | | |
| 5 | -467 | -383 | -439 | -467 | -467 | | |
| 6 | -1419 | -1329 | -1351 | -1419 | -1419 | | |
| 7 | -1403 | -1250 | -1321 | -1403 | -1403 | | |
| 8 | -956 | -735 | -825 | -957 | -956 | -738 | |
| 9 | -308 | -266 | -287 | -308 | -308 | | |
| 10 | -126 | -116 | -120 | -126 | -126 | | |
| 11 | -143 | -134 | -135 | -143 | -143 | | |
| 12 | -62 | -56 | -58 | -62 | -62 | | -58 |
| 13 | -136 | -117 | -124 | -136 | -136 | | |
| 14 | -260 | -215 | -240 | -260 | -260 | | -240 |
| 15 | -551 | -464 | -500 | -550 | -550 | | |
| 16 | -16 | -15 | -13 | -15 | -16 | -44 | |
| 17 | -129 | -111 | -119 | -130 | -129 | | |
| 18 | -62 | -56 | -59 | -62 | -63 | | |
| 19 | -129 | -116 | -119 | -129 | -129 | | |
| 20 | -281 | -231 | -251 | -281 | -281 | | |
| 21 | -50 | -48 | -47 | -51 | -50 | | |
| 22 | -300 | -262 | -282 | -300 | -300 | | |
| 23 | -280 | -227 | -246 | -280 | -280 | | |
| 24 | -110 | -97 | -103 | -110 | -110 | | |
| 25 | -365 | -307 | -341 | -365 | -365 | | |
| 26 | -1214 | -1073 | -1144 | -1214 | -1214 | -1096 | -1143 |

Table S4. Mean out-of-sample log-likelihoods for the communication app channel. The highest likelihoods are highlighted.

| Participant | Homogeneous | Hour (one-hot) | Hour (sine-cosine) | Day (one-hot) | Day (sine-cosine) | Hour + day (one-hot) | Hour + day (sine-cosine) |
|---|---|---|---|---|---|---|---|
| 1 | -694 | -616 | -655 | -694 | -694 | | |
| 2 | -938 | -779 | -840 | -937 | -937 | -770 | -839 |
| 3 | -1168 | -1136 | -1110 | -1168 | -1167 | -972 | -1109 |
| 4 | -116 | -109 | -112 | -116 | -116 | | |
| 5 | -417 | -347 | -393 | -417 | -417 | | -393 |

| | | | | | | | |
|---|---|---|---|---|---|---|---|
| 6 | -1268 | -1163 | -1193 | -1268 | -1268 | | |
| 7 | -1294 | -1136 | -1200 | -1293 | -1294 | -1141 | |
| 8 | -857 | -676 | -748 | -857 | -857 | -675 | -748 |
| 9 | -301 | -261 | -281 | -301 | -301 | | |
| 10 | -112 | -104 | -107 | -112 | -112 | | -107 |
| 11 | -126 | -118 | -118 | -126 | -126 | | |
| 12 | -60 | -54 | -57 | -60 | -60 | -54 | -57 |
| 13 | -127 | -111 | -116 | -127 | -127 | | |
| 14 | -239 | -201 | -221 | -238 | -238 | -200 | -220 |
| 15 | -514 | -438 | -471 | -514 | -514 | -438 | -470 |
| 16 | -14 | -14 | -13 | -13 | -14 | -46 | |
| 17 | -112 | -91 | -101 | -112 | -112 | | |
| 18 | -57 | -50 | -53 | -57 | -57 | | |
| 19 | -111 | -100 | -102 | -111 | -111 | | |
| 20 | -265 | -224 | -237 | -265 | -265 | | |
| 21 | -43 | -38 | -39 | -43 | -43 | | |
| 22 | -233 | -201 | -216 | -233 | -233 | -201 | -216 |
| 23 | -270 | -213 | -230 | -270 | -270 | | |
| 24 | -100 | -92 | -94 | -101 | -100 | | |
| 25 | -319 | -265 | -297 | -319 | -319 | -267 | |
| 26 | -1188 | -1051 | -1121 | -1187 | -1187 | -986 | -1121 |

Table S5. Mean out-of-sample log-likelihoods for the social media app channel. The highest likelihoods are highlighted.

| Participant | Homogeneous | Hour (one-hot) | Hour (sine-cosine) | Day (one-hot) | Day (sine-cosine) | Hour + day (one-hot) | Hour + day (sine-cosine) |
|---|---|---|---|---|---|---|---|
| 1 | -381 | -360 | -370 | -381 | -381 | -360 | -370 |
| 2 | -706 | -592 | -638 | -707 | -706 | | |
| 3 | -846 | -901 | -835 | -847 | -846 | -815 | -835 |
| 4 | -95 | -90 | -94 | -95 | -95 | | |
| 5 | -202 | -179 | -200 | -203 | -202 | -180 | -200 |
| 6 | -781 | -722 | -740 | -782 | -781 | -723 | -740 |
| 7 | -728 | -675 | -701 | -728 | -727 | | |
| 8 | -908 | -712 | -800 | -908 | -908 | -723 | |
| 9 | -168 | -157 | -162 | -168 | -168 | | |
| 10 | -89 | -84 | -88 | -90 | -90 | | |
| 11 | -92 | -90 | -88 | -92 | -92 | | |
| 12 | -37 | -35 | -36 | -38 | -37 | -36 | |
| 13 | -78 | -73 | -71 | -78 | -78 | | |
| 14 | -139 | -120 | -129 | -139 | -139 | -120 | -129 |
| 15 | -234 | -210 | -216 | -233 | -234 | -209 | -216 |
| 16 | -7 | -10 | -7 | -6 | -7 | -14 | |
| 17 | -109 | -91 | -100 | -109 | -109 | | |
| 18 | -22 | -27 | -21 | -23 | -22 | | -22 |
| 19 | -100 | -95 | -97 | -100 | -100 | -95 | -97 |

| | | | | | | | |
|---|---|---|---|---|---|---|---|
| 20 | -231 | -199 | -213 | -231 | -231 | | |
| 21 | -44 | -42 | -42 | -44 | -44 | | |
| 22 | -277 | -247 | -263 | -276 | -276 | -247 | -263 |
| 23 | -86 | -76 | -77 | -87 | -87 | -76 | -78 |
| 24 | -72 | -68 | -70 | -73 | -73 | | |
| 25 | -132 | -123 | -129 | -132 | -132 | | |
| 26 | -1010 | -924 | -983 | -1010 | -1011 | -924 | -984 |

Table S6. Multinomial logistic regression results from the total dwell time and age predictors, using the HMM that had imputation constrained based on phone usage.

| Total dwell time: | | | | | | |
|---|---|---|---|---|---|---|
| Group | Coefficient | Standard error | Odds ratio (95% CI) | *z* value | *P* value | FDR corrected *P* value |
| Schizophrenia | -0.0173 | 0.0158 | 0.9829 (0.9519-1.0138) | -1.0962 | 0.27 | 0.82 |
| Alzheimer's disease | -0.0530 | 0.0128 | 0.9484 (0.9232-0.9735) | -4.1369 | <.001 | <.001 |
| Subjective cognitive complaints | -0.0232 | 0.0080 | 0.9771 (0.9615-0.9926) | -2.9196 | 0.004 | 0.01 |
| Age: | | | | | | |
| Group | Coefficient | Standard error | Odds ratio (95% CI) | *z* value | *P* value | FDR corrected *P* value |
| Schizophrenia | -0.1493 | 0.0260 | 0.8613 (0.8105-0.9122) | -5.7521 | <.001 | <.001 |
| Alzheimer's disease | 0.0565 | 0.0265 | 1.0581 (1.0062-1.1101) | 2.1318 | 0.03 | 0.10 |
| Subjective cognitive complaints | 0.0034 | 0.0141 | 1.0034 (0.9757-1.0310) | 0.2405 | 0.81 | >.99 |

Table S7. Multinomial logistic regression results from the total dwell time and age predictors, using the HMM that had unconstrained imputation.

| Total dwell time: | | | | | | |
|---|---|---|---|---|---|---|
| Group | Coefficient | Standard error | Odds ratio (95% CI) | *z* value | *P* value | FDR corrected *P* value |
| Schizophrenia | -0.0176 | 0.0158 | 0.9826 (0.9516-1.0136) | -1.1120 | 0.27 | 0.80 |
| Alzheimer's disease | -0.0533 | 0.0129 | 0.9481 (0.9229-0.9733) | -4.1455 | <.001 | <.001 |
| Subjective cognitive complaints | -0.0234 | 0.0080 | 0.9769 (0.9613-0.9925) | -2.9398 | 0.003 | 0.01 |
| Age: | | | | | | |
| Group | Coefficient | Standard error | Odds ratio (95% CI) | *z* value | *P* value | FDR corrected *P* value |
| Schizophrenia | -0.1494 | 0.0260 | 0.8612 (0.8104-0.9121) | -5.7550 | <.001 | <.001 |
| Alzheimer's disease | 0.0563 | 0.0265 | 1.0579 (1.0059-1.1099) | 2.1205 | 0.03 | 0.10 |

| | | | | | | |
|---|---|---|---|---|---|---|
| Subjective cognitive complaints | 0.0033 | 0.0141 | 1.0033 (0.9757-1.0310) | 0.2341 | 0.81 | >.99 |
